# The Language & Memory Test: International feasibility study of an unsupervised digital test of cognition for neurologic populations

**DOI:** 10.1101/2025.01.28.25321275

**Authors:** Victoria M. Leavitt, Leila Simani, Marcus Koch, Sarah A. Morrow, Lauren Heuer, Mahrooz Roozbeh, Mehrdad Roozbeh, Sean Traynor

## Abstract

**Background:** Current cognitive outcomes are insufficient to monitor subtle change over time in neurologic populations. Challenging cognitive tests (i.e., cognitive “stress test”) unaffected by practice effects and learning are ideal for monitoring and tracking change. Here we describe the Language & Memory Test (LMT), an unsupervised digital test we developed to measure rapid naming, visuospatial memory, and fine motor dexterity, that takes approximately 4 minutes to complete.

**Objective:** Conduct feasibility study of the LMT in adults recruited across three countries; in English and non-English speaking participants; employing in-person, remote, and unsupervised administration modalities.

**Methods:** We investigated retest reliability, usability / tolerability, and equivalence across centers, administration modalities, and English / non-English speakers. We also evaluated performance across adult age groups: 18-40, 41-60, and 61+ years.

**Results:** The LMT was administered 555 times to 440 individuals ages 18-84 years in Canada, Iran, and the United States. Our sample was geographically and racially/ethnically diverse. Participants included a control sample of adults (n=115) and adults with a diagnosed neurologic disease (n=325). Of those invited to complete the LMT, 95.3% agreed; 99% of those who began the test completed it. Retest reliability was acceptable for finger tapping (Cronbach’s *α*=0.942) and language (*α*=0.872), and marginally acceptable for memory 1 (*α=*0.604). Equivalence varied across administration modalities, centers/countries, settings, and English / non-English speakers. Expected age-related performance decrements were shown for all three subtests.

**Conclusions:** The LMT shows promise as a culturally adaptable tool for use in diverse neurologic populations.

## INTRODUCTION

Cognitive decline is common in many neurologic diseases. Our ability to treat and understand cognitive impairment hinges on the availability of precise measurement tools to detect change over time. The need for appropriate cognitive outcomes in clinical trials has become more urgent since disease modifying therapies (DMTs) for several neurological diseases are being developed. One practical obstacle that stands in the way of successful cognitive outcomes is the time frame over which cognitive decline occurs. Clinical trials are designed to show an effect on a disease relevant measure within two or three years, an obvious mismatch with the remarkably slow pace of cognitive change in neurological diseases. In healthy adults over age 45, memory scores declined by -3.6% over a 10-year follow-up period, but drug trials generally incorporate follow-up periods that span much shorter time periods, e.g., 12-36 months.[1] Currently, clinical trials in neurologic populations often use classical instruments like the Montreal Cognitive Assessment (MoCA) and the Mini Mental Status Exam (MMSE). These and other instruments were developed as screening tests for dementia that capture cognitive status at a single timepoint; they were not developed to detect sometimes subtle changes over time.[2, 3] For neurologic populations, cognitive outcomes that can detect cognitive change from early disease stages, likely the best time to intervene in cognitive decline, is an unmet priority and need. Furthermore, there is a clear need for cognitive measurement tools that are relatively invulnerable to psychometric flaws like practice and learning effects, appropriate for clinical trial and clinical care workflow, and validated for unsupervised assessment. Ideally, such tools would be culturally adaptable, brief, inexpensive, and acceptable for both patients and clinicians from diverse populations.

We developed the Language & Memory Test (LMT), a digital unsupervised multimodal cognitive test, to optimize the detection of cognitive change over time in diverse neurologic populations. Leveraging digital technology, we focused on the following features based on practical, real-life demands of clinical care and research settings: brevity, tolerability, cultural adaptability, acceptability for individuals with low or no technology experience as well as diverse education levels, and feasibility for use in English and non-English speakers. Finally, and critically, the LMT incorporates tasks measuring cognitive domains most vulnerable to pathologic cognitive decline based on evidence from the cognitive neuroscience literature. [4–7] These include a rapid naming task involving spontaneous stimulus-bound retrieval, a function evidence suggests to be more sensitive to early deterioration than encoding tasks[6] and a function that is not measured by currently available unsupervised digital tools to our knowledge, and based on a recent scoping review.[8] The LMT additionally measures visuospatial memory, fine motor dexterity, and multitasking (as a latent variable).

This is a proof-of-concept feasibility study of the LMT. Our aim was to assess initial acceptability, tolerability, and psychometric properties of the LMT administered in geographically diverse sites, in English and non-English speaking individuals, in diverse neurologic populations, and utilizing diverse administration modalities (i.e., supervised and unsupervised, remote and in-person).

## METHODS

We followed the Strengthening the Reporting of Observational Studies in Epidemiology (STROBE) reporting guidelines, with the associated checklist for this study included as Supplementary Table 1.

### Study design

This is a cross-sectional observational study involving data collection at a single time point. A subsample of participants completed the LMT at a second time point to evaluate test-retest reliability. Data collection was initiated at each of three sites as local IRB approval was obtained, and took place from December 2023-November 2024.

### Participants

#### Sample size

Convenience sampling was employed to permit initial observations of how well tolerated the LMT would be for use in the context of a busy clinical setting and as a research tool. We therefore did not determine sample size a priori. Our use of convenience sampling allowed us to reach a large and diverse sample.

#### Inclusion/exclusion criteria

To be eligible, individuals were required to be over age 18-years of age. There were no exclusion criteria. Participants were patients and family members / friends of patients seen at in-person clinic visits at a general neurology practice (Tehran); patients and family members / friends of patients seen at an academic hospital (Calgary); research participants enrolled in a research study that included adults diagnosed with multiple sclerosis (MS) and a matched control group recruited through word-of-mouth (NYC-based remote study); and patients seen in-person at a general neurology clinic (NYC).

#### Demographic and clinical data

At enrollment, all participants provided their age and self-reported sex or gender. For individuals with neurologic conditions, diagnosis was confirmed via chart review.

#### Calgary cohort

Data collection took place at in-person clinic visits at the Calgary MS Clinic at the University of Calgary. Participants were enrolled and completed the LMT at one or two time points within the same clinic visit. The test was administered via iPad, in a face-to-face format with the clinician.

#### Tehran cohort

Data collection took place at in-person clinic visits at a general neurology clinic at Shahid Beheshti University. Participants were enrolled and completed the LMT at one time point within the clinic visit. The test was administered via iPad, in a face-to-face format with the clinician. Simple English instructions displayed in the app were translated orally for all participants by the administering clinician.

#### US-based remote cohort

Data collection took place via a remote video link-based research study for which baseline data collection was conducted from December 2023 to June 2024. Adults with MS and controls were enrolled from a total of 20 states within the continental US, Canada (5), the United Kingdom (3), France (1) and the Netherlands (1). Two time points of data were collected: first, participants were instructed to download the app to their personal device while completing the teleconference study visit (Time 1). Several days later (2-14 days), they received an email prompt from the study coordinator instructing them to self-administer the LMT (Time 2). A 3-item tolerability survey was administered to participants in this cohort: 1-Did you find the LMT to be tolerable; 2-Did you prefer the LMT to other tests; 3-Would you be willing to do the LMT daily or weekly for a research study.

### The LMT

The LMT was developed for iOS using SwiftUI, Python, and Flask app languages and frameworks. The LMT only exists in one version incorporating simple English-language instructions. For the site where English is not the primary language (Tehran), the test administrator (MR and MR) translated all instructions for the user in real time in the face-to-face setting. The LMT comprises 3 subtests. Total administration time is approximately 4 minutes. Tasks were developed to challenge cognitive functions most vulnerable to neurodegenerative disease-related changes,[4–7] while maximizing efficiency, tolerability, and acceptability for feasible and rapid uptake in clinical, research, and real-world settings. The on-screen presentation of LMT subtests is shown in **Figure 1**. Task 1 is a 10-second finger tapping task. Differences in speed of finger tapping are noted in neurodegenerative diseases even in the absence of primary motor symptoms.[9] In addition, this score can be used to control for motor confounds inherent to performance of all subsequent tests. Task 2 is a speeded retrieval test of lexical access using Language tasks (the ability of a participant to name a simple picture) and encoding (the ability of a participant to encode the position of a simple picture in a 3x3 or 4x4 grid). For each trial of the Language tasks, nine stimuli (pictures) are presented one at a time. Test taker must name the item pictured out loud as quickly as possible, then press the button. In the next screen, the item is presented for 3 seconds in a 3x3 grid. After nine items have been presented, the test taker is shown one of the pictures with a blank grid and must select the position on the grid where they saw the item before (Memory 1). After each response, feedback is provided (selected grid quadrant green=correct, selected grid quadrant=incorrect). Once again, the same nine items are presented one at a time and the test taker must name each item as quickly as possible and press the button (Language 2). In the following screen, the item is presented for 3 seconds on a 4x4 grid. When all nine items have been presented twice, Memory 2 commences. See **Figure 1**.

**Figure 1.**
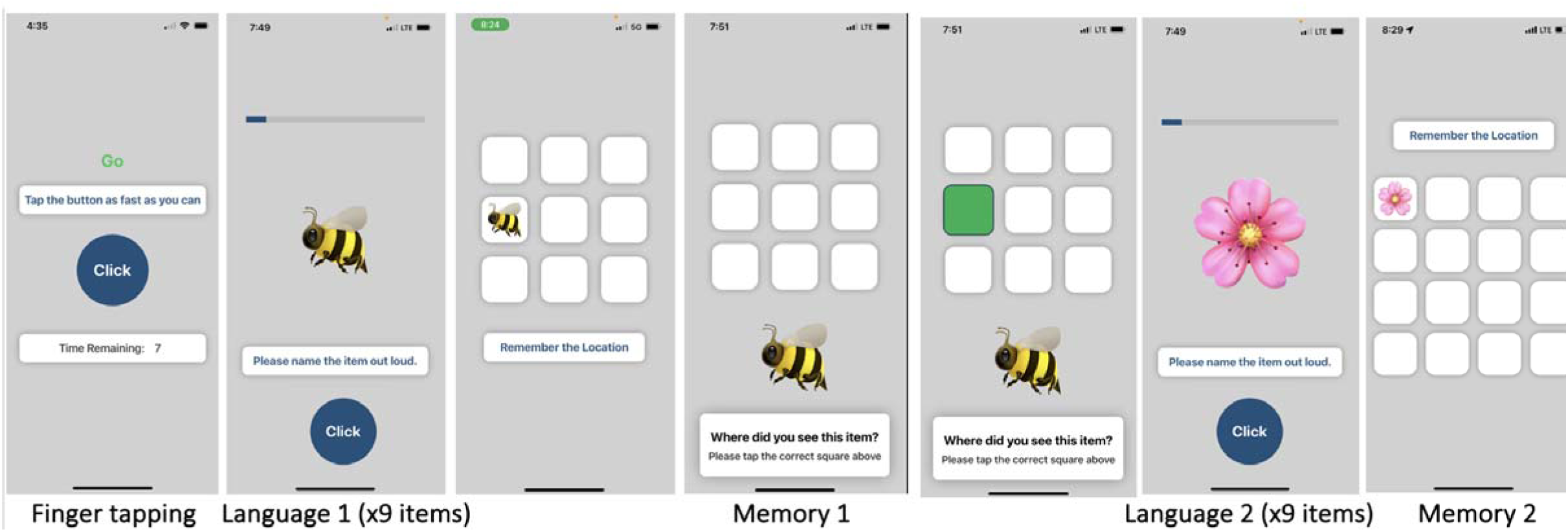
LMT subtests: Finger tapping, Language 1 and 2, Memory 1 and 2.

#### Analytic approach

Demographic and clinical characteristics of participants were evaluated within and between groups using parametric and non-parametric statistics. All t-tests were two-sided. Significance threshold was set at *p*< .05. For the Language subtest, the mean of Language trial 1 and Language trial 2 was calculated. This mean Language score was further regression-adjusted to account for the confound of fine motor speed as follows: a regression-adjusted score was calculated to account for the confounding influence of fine motor speed by entering Finger tapping score as the independent variable and Language mean score as the dependent variable. This regression-adjusted score was used in all analyses in which between-group comparisons of subtest performance were made.

Reliability was evaluated two ways: first, a metric of internal consistency was calculated using scores for the two Language trials. In both trials, the same nine items are presented, although not in the same order. Therefore, it is not a perfect test of internal consistency since we would reasonably expect participants to retrieve the word faster the second time it is presented. Nonetheless, we evaluated concordance across trials as a proof-of-concept check of internal consistency by calculating Cronbach’s alpha (*α*). Second, test-retest reliability was evaluated in the subsample of participants who completed the LMT twice. This included participants in the US-based research sample who completed the LMT the first time in a supervised remote video-link enabled study visit and the second time via unsupervised self-administration on their personal smartphone 1-2 weeks later (after receiving a reminder from study personnel). In the Calgary sample, participants were administered the LMT twice within the same clinical visit. Cronbach’s alpha was calculated as an estimate of retest reliability for Time 1 and Time 2 scores for all subtests.

For analysis of age-based differences, the full sample (across all three centers) was separated into the following age windows: 18-40, 41-60, 61+ years. Analysis of covariance was used to compare performance on all subtests across age windows first in the full sample, followed by separately within the Clinical and Control subsamples.

For equivalence testing, group differences were evaluated using analysis of covariance controlling for variables that differed between groups.

### Standard Protocol Approvals, Registrations, and Patient Consents

This study involving human participants was performed in line with the principles of the Declaration of Helsinki. The study was approved by local institutional review boards at each participating site. Written informed consent was obtained from all participants at time of enrollment.

### Data Availability

The dataset is available from the corresponding author upon reasonable request.

## RESULTS

Data collection was begun in the US-based remote cohort in December 2023, followed by Tehran in January 2024, and Calgary in February 2024. Data collection was finalized on November 30, 2024. A total of 440 participants were enrolled: US-based cohort (N=89), Tehran (N=185), and Calgary (N=166); see **Table 1**.

**Table 1.** Sample characteristics.

|  | **Overall (N=440)** | **Clinical (N=325)** | **Controls (N=115)** | ***p*-value** |
| --- | --- | --- | --- | --- |
| Center |  |  |  |  |
| NYC / Tehran / Calgary | 89 / 185 / 166 | 51 / 152 / 122 | 38 / 33 / 44 | - |
| Sex, Women (%) | 324 (73.6) | 254 (78.2) | 70 (60.9) | <0.001 |
| Mean age, years (SD) | 44.1 (13.1) | 45.3 (12.6) | 40.5 (13.8) | <0.001 |
| Age range | 18-84 | 18-84 | 18-83 | - |
| Race / ethnicity, N (%) |  |  |  | 0.205 |
| White | 408 (92.7) | 302 (92.9) | 106 (92.2) |  |
| Black | 18 (4.1) | 15 (4.6) | 3 (2.6) |  |
| Asian | 14 (3.2) | 8 (2.5) | 6 (5.2) |  |
| Hispanic | 6 (1.4) | 3 (1.0) | 3 (2.6) |  |
| <u>Neurologic population</u>, N (%) |  |  |  |  |
| Multiple sclerosis | - | 265 (60.2) | - | - |
| Other |  | 60 (18.5) |  |  |

### US-based cohort

For the US-based cohort, English-speaking adults with MS (n=61) and controls (n=21) were enrolled in a research study via personal email invitation from 20 states within the continental US (n=72), Canada (n=5), the United Kingdom (n=3), France (n=1) and the Netherlands (n=1). Additional participants were enrolled in the context of a clinical visit at Columbia University’s Department of Neurology (n=7). Research participants took part in a video-link enabled 1-hour study visit during which they downloaded the LMT to their personal smartphone. They were then requested to repeat the LMT one to two weeks later in an unsupervised administration; of these, 82.6% complied.

### Tehran cohort

Participants enrolled in the Tehran cohort were neurology clinic patients seen in-person at an academic medical center, and recruited by their clinician. The cohort included 92 patients with MS, 60 patients with other neurologic conditions [MCI/dementia (16), migraine (22), Parkinson’s disease (2), stroke (2), epilepsy (1), depression (5), and neuromuscular disorders (12)], and 44 controls.

### Calgary cohort

The Calgary cohort was comprised of individuals seen in the context of standard clinical care at an academic MS center; based on direct invitation from their clinician, 122 patients with MS and 44 controls were enrolled.

Sample characteristics for participants across the three centers are displayed in **Table 1**. The majority of clinical patients were diagnosed with multiple sclerosis (MS), with additional neurologic patients being seen for cognitive issues secondary to neurologic conditions (e.g., dementia, migraine, Parkinson’s disease, etc.).

Acceptability of the LMT was high: of those invited to complete the test, 95.3% agreed. Of those who declined, 14 (enrolled in the remote cohort) were unable to download the task because they had an Android phone, 5 (seen in clinic visits: 1 in Calgary, 4 in Tehran) did not have time, and 1 (remote cohort) was not comfortable with technology. Usability of the test was high: 99.3% of individuals who began the test completed it. Two participants failed to complete the first memory subtest, Memory 1, and an additional two stopped on the final, most difficult memory test, Memory 2.

### LMT Subtest Performance

Mean scores (SD) are displayed in **Table 2** for each of the three subtests for the full sample, Clinical patients, and Controls, respectively. On each task and as expected, the Clinical sample performed consistently worse than the Control group even after controlling for age and sex/gender differences across groups (*p*’s from .025 to <.001).

**Table 2.** LMT subtest performance.

|  | Overall (N=440) | Clinical (N=325) | Control (N=115) | *p*-value | *p*-value adj* |
| --- | --- | --- | --- | --- | --- |
| Finger tapping<br>Total taps (mean, SD) | 55.0 (14.2) | 52.6 (14.0) | 61.8 (12.3) | <0.001 | <0.001 |
| Language, seconds<br>Total seconds (mean, SD) | 26.4 (10.4) | 27.8 (11.0) | 22.1 (6.6) | <0.001 | <0.001 |
| Memory 1, % accurate<br>(mean, SD) | 52.8 (26.5) | 50.1 (26.1) | 60.4 (26.3) | <0.001 | <0.001 |
| Memory 2, % accurate<br>(mean, SD) | 36.2 (21.4) | 34.6 (20.1) | 40.9 (24.3) | 0.007 | 0.025 |

\*Adjusted for age and sex/gender.

For the language task, as noted, we calculated the average of two interleaved trials and used this single metric in all subsequent analyses except the reliability analyses. We also inspected performance on each respective Language trial and found that the full group improved in their ability to rapidly name the nine items from trial one (mean seconds 27.7 ± 12.1) to trial two (25.0 ± 10.8); t=48.0, *p<*.001. This difference was maintained for both the Clinical group (Trial 1: 29.4 ± 12.7, Trial 2: 26.2 ± 11.5) and the Controls (Trial 1: 22.7 ± 8.4, Trial 2: 21.6 ± 7.4); both *p*’s*<*.001.

### Reliability and internal consistency of LMT subtests

Test-retest reliability was estimated by calculating Cronbach’s alpha (*α*) for each of the three subtests; see **Table 3a**. In the full sample, Finger tapping and Language demonstrated high reliability. Whereas Memory 1 showed marginal acceptability (*α*=.604), the reliability of Memory 2 was unacceptable (*α*=.418). Within the Clinical group, Finger tapping and Language demonstrated high reliability, but Memory 1 (*α*=.479) and Memory 2 (*α*=.293) fell below the generally accepted threshold of 0.7. In the Control group, only Memory 2 demonstrated low reliability, *α*=.589. Next, as a test of internal consistency, we evaluated performance across the two trials of the language task within a single administration of the LMT in the full sample. For the full sample as well as within the Clinical and Control subsamples, reliability was acceptable (*α=*.872 overall). Full results are presented in **Table 3b**.

**Table 3.** Test-retest reliability (3a) and internal consistency (3b) as estimated by Cronbach’s alpha.

| **3a. Participants who took the LMT at two time points.** |  |  |  |
| --- | --- | --- | --- |
|  | Overall (N=192) | Clinical (N=136) | Controls (N=56) |
| Finger tapping | .942 | .951 | .880 |
| Language | .872 | .898 | .691 |
| Memory 1 | .604 | .479 | .774 |
| Memory 2 | .418 | .293 | .589 |
| **3b. Measures of internal consistency for the full sample.** |  |  |  |
|  | Overall (N=440) | Clinical (N=325) | Controls (N=115) |
| Language 1 and Language 2 | .872 | .898 | .691 |

### Age-based subtest performance differences across the adult lifespan

To evaluate expected subtest performance differences across the adult lifespan (i.e., steadily declining cognitive performance), we separated the group into young adult (age 18-40), middle adult (41-60) and older adults (61-85), and analyzed the scores for each subtest. In **Table 4** and **Figure 2**, we show that the expected gradient of decline over the adult life course for each of the subtests in the full group. We next evaluated these differences separately in the clinical and control groups. Differences were significant for all tasks in the clinical group; in the controls, differences were significant for Finger tapping and Language tasks (*p<*.001). For the memory tasks, however, differences did not reach the level of significance (Memory 1: *p=*.26, Memory 2: *p=*.08). Age-based performance differences are displayed for each group separately in **Table 5**.

**Figure 2.**
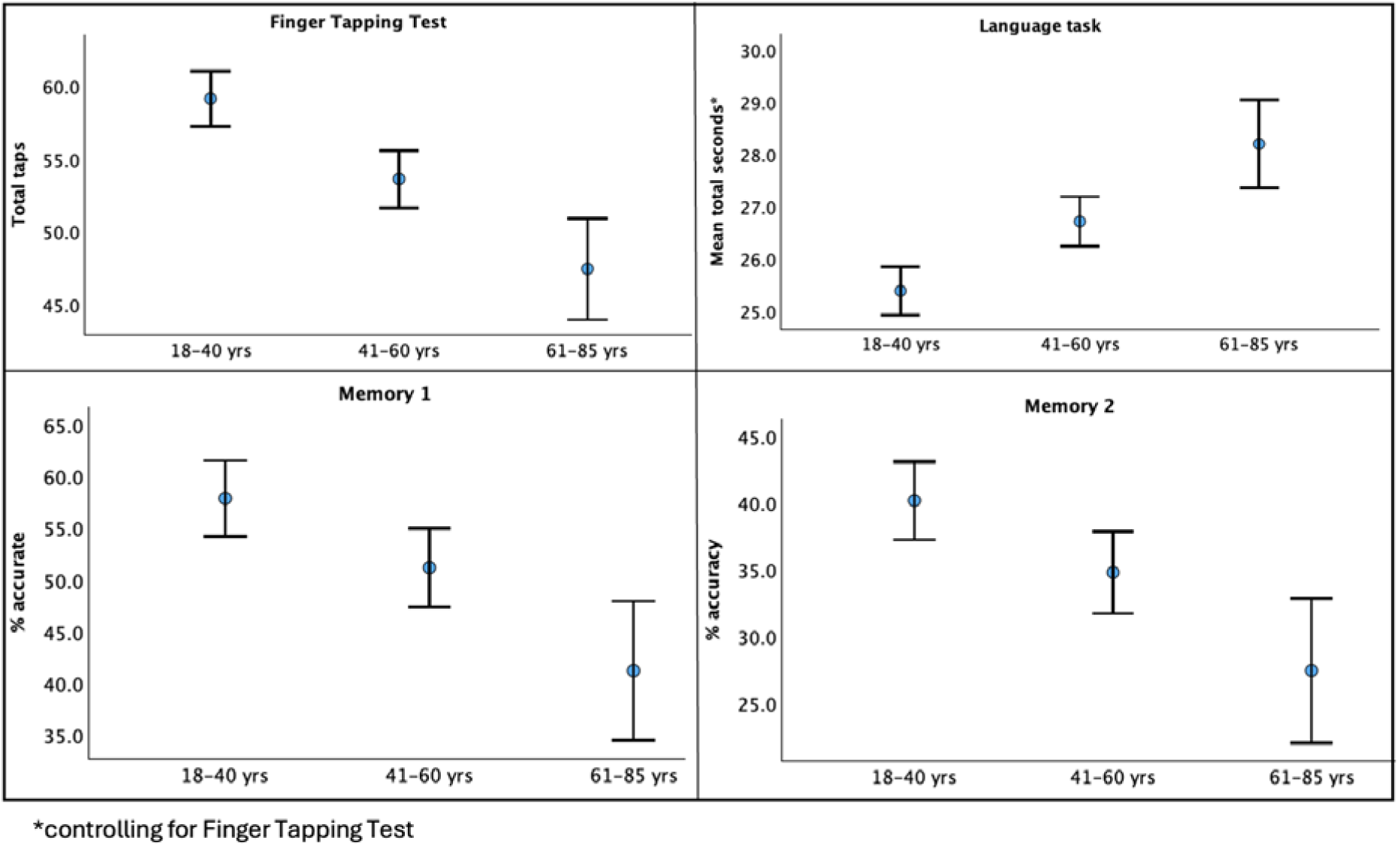
Age based subtest performance. Differences across age windows significant for all subtests, *p*<.001.

**Table 4.** Age-based differences in performance on each subtest (N=436). In the full group, differences across age windows were significant for all tasks (*p<*.001).

|  | Age window | Mean | SD | N |
| --- | --- | --- | --- | --- |
| Finger Tapping Test (FTT) | 18-40 | 59.12 | 12.23 | 196 |
|  | 41-60 | 53.61 | 14.41 | 182 |
|  | 61-85 | 47.45 | 14.50 | 58 |
|  | Total | 55.27 | 14.03 | 436 |
| Language (controlling for FTT) | 18-40 | 25.37 | 2.95 | 196 |
|  | 41-60 | 26.70 | 3.47 | 182 |
|  | 61-85 | 28.19 | 3.50 | 58 |
|  | Total | 26.30 | 3.39 | 436 |
| Memory 1 | 18-40 | 57.84 | 26.81 | 196 |
|  | 41-60 | 51.15 | 26.38 | 182 |
|  | 61-85 | 41.19 | 22.32 | 58 |
|  | Total | 52.83 | 26.60 | 436 |
| Memory 2 | 18-40 | 40.14 | 21.91 | 196 |
|  | 41-60 | 34.75 | 21.05 | 182 |
|  | 61-85 | 27.39 | 17.93 | 58 |
|  | Total | 36.19 | 21.45 | 436 |

**Table 5.** Age-based differences in performance on each subtest within each group. All differences are significant in the clinical Group (*p*<.001); in the controls, Finger Tapping and Language were significant at *p*<.001. Memory task differences were not significant.

| Controls |  |  |  |  | Clinical |  |  |  |  |
| --- | --- | --- | --- | --- | --- | --- | --- | --- | --- |
|  | Age Window | Mean | SD | N |  | Age Window | Mean | SD | N |
| Finger Tapping Test (FTT) | 18-40 | 65.24 | 9.72 | 68 | Finger Tapping Test | 18-40 | 55.88 | 12.22 | 128 |
|  | 41-60 | 62.00 | 8.57 | 31 |  | 41-60 | 51.89 | 14.77 | 151 |
|  | 61-85 | 50.07 | 15.78 | 14 |  | 61-85 | 46.61 | 14.15 | 44 |
|  | Total | 62.47 | 11.36 | 113 |  | Total | 52.75 | 14.02 | 323 |
| Language (controlling for FTT) | 18-40 | 23.90 | 2.34 | 68 | Language (controlling for FTT) | 18-40 | 26.16 | 2.95 | 128 |
|  | 41-60 | 24.68 | 2.07 | 31 |  | 41-60 | 27.12 | 3.56 | 151 |
|  | 61-85 | 27.56 | 3.81 | 14 |  | 61-85 | 28.39 | 3.41 | 44 |
|  | Total | 24.57 | 2.74 | 113 |  | Total | 26.91 | 3.38 | 323 |
| Memory 1 | 18-40 | 62.78 | 28.42 | 68 | Memory 1 | 18-40 | 55.21 | 25.63 | 128 |
|  | 41-60 | 60.57 | 23.98 | 31 |  | 41-60 | 49.21 | 26.51 | 151 |
|  | 61-85 | 50.00 | 19.86 | 14 |  | 61-85 | 38.38 | 22.54 | 44 |
|  | Total | 60.59 | 26.46 | 113 |  | Total | 50.11 | 26.15 | 323 |
| Memory 2 | 18-40 | 44.46 | 24.57 | 68 | Memory 2 | 18-40 | 37.85 | 20.08 | 128 |
|  | 41-60 | 38.35 | 23.63 | 31 |  | 41-60 | 34.01 | 20.49 | 151 |
|  | 61-85 | 29.36 | 21.62 | 14 |  | 61-85 | 26.77 | 16.83 | 44 |
|  | Total | 40.91 | 24.31 | 113 |  | Total | 34.54 | 20.13 | 323 |

### Equivalence across center / country

We were interested in determining equivalence across center / country. Ideally, performance of test-takers should be roughly equivalent regardless of the country in which the LMT is being taken. We first evaluated differences in sex and age across centers, finding significant differences (*p<*.001) for sex / gender (see **Table 6)**, which we controlled for in subsequent analyses. Based on analysis of covariance, we found center differences in performance on all subtests (*p<*.001, **Table 7**). Effect sizes for differences were small (η_p_^2^: .069 - .177). Separating the sample by group (clinical and control), these differences were maintained for all subtests except Memory 1 in the Control group.

**Table 6.**
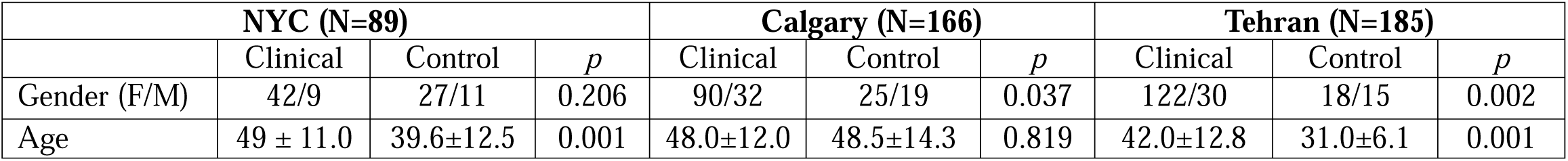
Sample characteristics based on center.

| **NYC (N=89)** |  |  |  | **Calgary (N=166)** |  |  | **Tehran (N=185)** |  |  |
| --- | --- | --- | --- | --- | --- | --- | --- | --- | --- |
|  | Clinical | Control | *p* | Clinical | Control | *p* | Clinical | Control | *p* |
| Gender (F/M) | 42/9 | 27/11 | 0.206 | 90/32 | 25/19 | 0.037 | 122/30 | 18/15 | 0.002 |
| Age | 49 ± 11.0 | 39.6±12.5 | 0.001 | 48.0±12.0 | 48.5±14.3 | 0.819 | 42.0±12.8 | 31.0±6.1 | 0.001 |

**Table 7.** LMT subtest scores by center, controlling for age.

| | NYC N=89 | Calgary N=166 | Tehran N=185 | Adjusted *p* | $\eta_p^2$ |
| --- | --- | --- | --- | --- | --- |
| **Overall** |  |  |  |  |  |
| Finger tapping | 54.6 ± 13.2 | 57.5 ± 12.6 | 53.1 ± 15.7 | <.001 | .177 |
| Language | 23.5 ± 6.6 | 26.5 ± 8.4 | 27.4 ± 12.9 | <.001 | .146 |
| Memory 1 | 54.8 ± 23.9 | 55.7 ± 26.6 | 49.3 ± 27.3 | <.001 | .079 |
| Memory 2 | 40.0 ± 22.1 | 35.9 ± 19.3 | 34.7 ± 22.7 | <.001 | .069 |
| **Clinical N=323** |  |  |  |  |  |
| Finger tapping | 49.9 ± 14.4 | 57.3 ± 11.0 | 49.8 ± 15.1 | <.001 | .192 |
| Language | 24.3 ± 6.7 | 27.5 ± 8.6 | 29.1 ± 13.4 | <.001 | .155 |
| Memory 1 | 48.4 ± 22.0 | 55.0 ± 26.3 | 46.7 ± 26.8 | <.001 | .082 |
| Memory 2 | 35.3 ± 18.3 | 36.7 ± 19.3 | 32.5 ± 21.3 | <.001 | .056 |
| **Controls N=115** |  |  |  |  |  |
| Finger tapping | 60.8 ± 8.0 | 58.0 ± 16.3 | 67.9 ± 7.3 | <.001 | .159 |
| Language | 25.0 ± 1.9 | 25.6 ± 3.9 | 23.2 ± 1.8 | .018 | .088 |
| Memory 1 | 63.2 ± 24.4 | 57.9 ± 28.1 | 60.9 ± 27.0 | .394 | .027 |
| Memory 2 | 46.2 ± 25.1 | 33.6 ± 19.6 | 44.1 ± 27.1 | .018 | .088 |

### Equivalence across mode of administration: In-person, remote, unsupervised

Comparing in-person versus remote supervised administration, we found no significant differences (see **Table 8a)**. We next evaluated differences between the small subset of participants who completed the LMT in an unsupervised setting (N=66), US-based cohort participants who downloaded the LMT onto their personal smartphone and self-administered it at a second time point. Comparing this subsample to the rest of the sample who completed the LMT at a second time point in a supervised setting (N=125) revealed small effect size differences on the Finger Tapping task (p=.032, η_p_^2^ = .024) and Memory 1 (p=.012, η_p_^2^ = .033) in the overall sample (**Table 8b**).

**Table 8.** LMT subtest performance by mode of administration: (8a) in-person versus remote supervised, and (8b) unsupervised versus supervised (data from visit 2 only).

| **8a. In-person versus remote supervised administration.** |  |  |  |
| --- | --- | --- | --- |
|  | **In-person**<br>N=370 | **Remote**<br>N=66 | *p*-value |
| Finger tapping | 55.4 ± 14.3 | 54.7 ± 13.2 | .443 |
| Language | 26.3 ± 3.4 | 26.4 ± 3.2 | .061 |
| Memory 1 | 52.3 ± 27.2 | 54.8 ± 24.1 | .689 |
| Memory 2 | 35.2 ± 21.1 | 40.0 ± 22.1 | .111 |
| **8b. Unsupervised versus supervised administration. Effect size ($\eta_p^2$) is provided for significant *p*-values.** |  |  |  |
| | **Unsupervised**<br>N=66 | **Supervised**<br>N=125 | *p*-value, $\eta_p^2$ |
| Finger tapping | 56.0 ± 14.4 | 59.9 ± 10.4 | .032, .024 |
| Language | 22.9 ± 6.3 | 24.4 ± 7.7 | .179 |
| Memory 1 | 64.8 ± 25.2 | 55.1 ± 24.9 | .012, .033 |
| Memory 2 | 35.2 ± 21.0 | 35.0 ± 19.2 | .957 |

### Equivalence in English and non-English speakers

Equivalence of LMT subtest performance in English and non-English speakers was evaluated. Here, all tasks showed small effect size differences in the overall sample (η_p_^2^: .066-.165); see **Table 9**.

**Table 9.** LMT subtest performance by English versus non-English speakers.

| | **English** | **Non-English** | *p*-value | $\eta_p^2$ |
| --- | --- | --- | --- | --- |
| **Overall** |  |  |  |  |
| Finger tapping | 56.5 ± 12.9 | 53.1 ± 15.7 | <.001 | .165 |
| Language | 25.5 ± 7.9 | 27.5 ± 13.0 | <.001 | .141 |
| Memory 1 | 55.4 ± 25.6 | 49.3 ± 27.3 | <.001 | .076 |
| Memory 2 | 37.3 ± 20.4 | 34.7 ± 22.7 | <.001 | .066 |

### Tolerability and usability

A 3-item tolerability survey administered to the US-based cohort participants (N=66) revealed acceptable tolerability: 100% reported the LMT to be tolerable, 91% reported that they would be willing to complete the LMT daily for a research study, if requested; 18% preferred the LMT to classic paper-and-pencil neuropsychological test measures. Across all three centers, no participants were unable to complete the task due to technology demands.

### Data availability

Data used for this study are available upon request to the corresponding author.

## DISCUSSION

This proof-of-concept, multinational feasibility study supports the LMT, a brief, multimodal digital cognitive test designed to be an unsupervised tool, as accessible and acceptable for measuring cognition in diverse neurologic populations. Our study was conducted across three countries on two continents, enrolling participants two ways: recruited for a research study in the course of standard in-person clinical care, and recruited into a home-based remote research study. We tested the LMT using tablet-based as well as smartphone administration, through in-person administration with a tester, via video conference, and in an unsupervised format. Finally, we enrolled racially and ethnically diverse participants, and included English and non-English speaking individuals across the adult lifespan.

### The rapidly evolving landscape of unsupervised digital tools

Digital tools for measuring cognition are rapidly being developed to meet the demand for reliable, accurate, inexpensive measurement tools leveraging technology to optimize sensitivity to, e.g., biomarkers for preclinical Alzheimer’s disease (AD),[10–12] and subtle patient-reported cognitive changes in neurologic conditions affecting young adults such as multiple sclerosis (MS).[13] Such tools are urgently needed for research and clinical use in neurologic populations. A recent scoping review identified 23 unsupervised digital assessment tools that have been developed for detection of preclinical AD.[8] Several features distinguish the LMT: (1) Incorporation of a speeded task of retrieval using a simple picture naming task. Picture naming tasks (e.g., the Boston Naming Task) have been an indispensable component of classic neuropsychological batteries used for diagnosing neurodegenerative conditions such as AD. Growing evidence suggests that the breakdown of stimulus-dependent spontaneous retrieval may be an early sign of abnormal cognitive aging.[6, 14] We leveraged this vulnerability to create a more challenging and speeded version of a task incorporating confrontation naming, rapid lexical access, and spontaneous stimulus-bound retrieval. (2) In contrast to available tools, many of which involve batteries of as many as 23 tests and often include word list learning tasks not readily adaptable for use in non-English speakers, the LMT is a brief, all-in-one test combining function across four cognitive domains and can be administered to non-English speaking individuals. (3) Acceptable tolerability / usability is critical to the adoption and successful uptake of a tool for clinical and research use.

### Metadata

We designed the LMT to incorporate multiple manipulable features that yield meta-data beyond the three subtask scores. These meta-data can be used to inform a number of ancillary hypothesis-driven questions. First, the LMT includes timed and untimed tasks that can be used to isolate the impact of speed on task performance. Second, the LMT incorporates an expressive speech component, enabling us to capture and analyze speech features such as time to initiation and speech articulation for future research. Third, the LMT provides performance feedback during the memory task: after each response, the square in the grid either turns green (correct) or red (incorrect). Use of feedback to improve performance is a latent indicator of learning capacity, which holds discriminative value for some neurologic populations.[15] Finally, several parameters of the memory task could be used to test specific hypotheses in future research. For instance, inspection of item-level data would permit interrogation of the object- versus spatial-based hypothesis of AD: memory for object-based (anterior-temporal) versus spatial-based (posterior-medial) information[16] is differentially affected by tau and amyloid-ß deposition.[17, 18] Differences in stimuli position, i.e., upper versus lower visual field, could be used to test age-related spatial mnemonic deficits.[19] That is, memory for everyday objects presented in upper versus lower visual fields is shown to be worse for older adults compared to younger adults, a distinction that could be diagnostically useful. These and other hypotheses could be directly testing using available meta-data from the LMT.

### A latent test of multitasking

The LMT incorporates a challenging latent test of multitasking. Because of the interleaved design, the LMT possesses a latent multitasking demand that remains to be quantified in future work through extraction of precise timestamps to demarcate each individual subtest. That is, the test in its current form yields a single score for the language task that combines the total time (in seconds) for a participant to name an item, advance (via button press) to the next screen, and then view and remember (encode) the item in its location displayed in a grid. Only one of these constituent parts (name the item) is the actual lexical retrieval task, but all three together constitute a challenging test of multitasking. Extracting a score that isolates multitasking remains to be accomplished in future work, but it is notable that to date, few (if any) widely accepted neuropsychological tests of multitasking exist.

### Limitations / Opportunities for improvement

Based on this initial proof-of-concept study, we identified unacceptable psychometric properties of the Memory tasks, which failed to demonstrate strong test-retest reliability: with our sample demonstrating mean accuracy of 36.2% (Memory 2), the task in its current state is too difficult for most participants regardless of diagnostic group. To address this, we will incorporate a modification to the next iteration of the LMT: challenge levels that can be pre-selected and matched to specific demographic, clinical, and/or performance metrics to tailor the task to each individual. The failure to find evidence supporting equivalence across centers in the clinical group is a cause for concern that warrants further study in future work. Interestingly, cultural differences in test-taking factors including motivation have been noted in prior research (including the observation that getting a high score on a cognitive test is not a universal value) and merit investigation in future studies.[20] The high level of acceptability, usability, and tolerability shown consistently across centers is encouraging. Finally, although there was a lack of racial and ethnic diversity of the full sample, the US-based sample was 18% Black, 6.7% Asian, and 6.7% Hispanic. In a multinational study including countries with low racial/ethnic heterogeneity, our sample composition is not unexpected.

In summary, the LMT shows promise as a useful unsupervised tool for measuring cognitive change in diverse neurologic populations to potentially serve as a clinical trial outcome and as a candidate cognitive stress test to advance a basic understanding of cognitive capacity over the life course, across cultures, and in individuals who historically have been underrepresented in research studies. The LMT is brief: at approximately 4 minutes to complete, it is one of the shortest multimodal tests available. The results of this initial feasibility study suggest the LMT’s acceptability as a cognitive outcome that can be seamlessly integrated into clinical care and research.

## Data Availability

All data produced in the present study are available upon reasonable request to the corresponding author (VML).

## ACKNOWLEDGMENTS

The authors wish to thank the many individuals who participated in this research study. We also thank Jason Hassenstab, Martin Sliwinski, Laura Germine, and Adam Staffaroni for feedback early in the development of the LMT prototype.

